# Reflection of connectivism in medical education and learning motivation during COVID-19

**DOI:** 10.1101/2020.07.07.20147918

**Authors:** Jun Xin Lee, Ahmad Hathim Ahmad Azman, Jing Yi Ng, Noor Akmal Shareela Ismail

**Author notes:** These authors contributed equally to this work (Jun Xin Lee, Ahmad Hathim Ahmad Azman, Jing Yi Ng).

## Abstract

The COVID-19 pandemic has not only affected the global healthcare and economy but threatened the world of education altogether. Malaysia is not spared from this pandemic as all universities were forced to close and initiate online learning with the implementation of Movement Control Order since mid-March 2020.The abrupt shift from conventional medical education to fully virtual learning definitely deserves a reflection on how it affects the learning motivation among medical students. Hence, this is the first study that compares the effect of digital learning on learning motivation among medical students in Universiti Kebangsaan Malaysia (UKM) prior to and during the COVID-19 pandemic. A modified Students Motivation towards Science Learning (SMTSL) was used to assess the learning motivation of UKM medical students throughout Year 1-5. The number of students that use digital learning during COVID-19 is significantly higher compared to before COVID-19 (p<0.05). However, there is no significant difference (p=0.872) in learning motivation among medical students before and during COVID-19 crisis. Higher frequency in digital learning usage frequency does not exert a great impact on learning motivation. Reflections from each participant were collated to justify the current situation. This could be due to motivation coming from the very choice to pursue medicine as a doctor, which is mainly influenced by intrinsic motivation, and ability to adapt in difficult situations. Thus, medical educators should be creative in enhancing extrinsic motivation by making use of digital learning as a platform so that medical students are able to independently fish for information in the vast pool of digital information and apply in actual medical practice in the future for life-long learning.

## Introduction

The sudden emergence of novel Coronavirus Disease 2019 (COVID-19) in the city of Wuhan, China, which subsequently spread to the entire world almost overnight has put the global public health under emergency with declaration of global pandemic by the World Health Organization. The sudden upsurge of cases due to a religious gathering with the nation’s first two deaths has put Malaysia under the implementation of nationwide Movement Control Order (MCO) with the hope to reduce community spread and avoid over burning the nation’s health system as much of the virus remains unknown and obscured [1]. Fortunately, the 3-month-MCO has shown promising success with steady reduction of local cases and introduction of recovery MCO which is part of the exit strategy that allows resumption of most economic sectors. Despite Malaysia is currently under recovery stage, there is no doubt that the COVID-19 pandemic has not only affected the global health and economy, but also the world of education. Up to 140 country-wide closures of schools and universities affecting more than 1 billion students were reported by the United Nations Education, Scientific and Cultural Organization (UNESCO). There is no exception to Malaysia as all learning is forced to move onto online space since the commencement of MCO. Regardless of the reopening of economic sectors, the Ministry of Higher Education Malaysia has announced that online learning will be continued in all universities until 31st of December 2020. The sudden shift away from conventional medical education into solely virtual learning possesses a great challenge for both medical educators and medical students as medicine is well-known to be an evidence-based field that involves internships or clerkships to learn from real patients [2]. Apart from that, issues concerning internet connection and unavailability of clinical equipment among medical students further increase the pressure and challenge in delivering medical education. Nevertheless, looking at the bright side, the COVID-19 pandemic has actually created a platform for medical educators to be more creative and innovative, exploring new learning designs or pedagogies to provide medical students with the best possible learning experiences [3]. Learning has also become more student-centered, encouraging self-directed learning with no time and place restrictions.

Indeed, digital learning has been incorporated into medical education since before the COVID-19 outbreak with production of medical doctors using the state-of-the-art technologies via the emergence of virtual universities overseas [4]. In Malaysia, the introduction of the Learning Management System (LMS) in conjunction with the ministry’s initiative, MyHE4.0 (Education 4.0) via the Higher Education Blueprint 2015-2025 has created a ‘blended learning’ environment for the students to communicate virtually with lecturers, submit assignments, carry out discussions and group work [5]. Studies have shown that most medical students are tech savvy and they support the use of digital learning in medical education as it is interactive, cost-effective, making the learning contents readily available [6-8]. However, students and educators are pressured with increasing time constraints and demands [9], together with network issues, availability of infrastructures and perception of being distracted while using high end technologies [6] have put digital learning only as a complementary role in medical education. With the abrupt introduction of fully online learning in medical education, it remains unknown regarding the trend of usage and acceptance of this new method of learning among medical students as compared to prior COVID-19 crisis period.

Over the years, research on the outcome of digital learning in medical education has been focusing on the level of confidence [10-12] and academic achievements or performance [10, 13, 14], which shows a positive relationship. Studies have also found a positive association between digital learning and learning motivation, but the respondents were all comprising undergraduates of courses other than medicine [15, 16]. Recently, there are no studies investigating the relationship between digital learning and learning motivation specifically among medical students, which is a vital aspect as learning motivation especially autonomous motivation is associated with better outcome of learning [17, 18], resilience [17] and thus motivation for lifelong learning [17-19].

Hence, it is crucial to explore the impact of solely digital learning on learning motivation among medical students during the current setting and to see whether there is any difference compared to before COVID-19 outbreak period in which students are still being taught with the usual conventional approaches of medical education. Besides, it would be essential to explore the effect of fully digital learning on connectivism which is a relatively new learning theory in this digital age among medical students. Thus, this study aims to identify the preferred sources of digital learning, investigate the difference in frequency of digital learning usage and difference in learning motivation, effect of digital learning on learning motivation among medical students of National University Malaysia (Universiti Kebangsan Malaysia, UKM) before and during COVID-19 pandemic period.

## Methodology

### Study Design

This was a cross sectional study that uses a quantitative approach, involving undergraduate medical students of Universiti Kebangsaan Malaysia (UKM). All UKM students undertaking the course of Doctor of Medicine during the period of this study were eligible for this study. Students from other courses, other universities, and who did not understand English were all excluded. Stratified convenience sampling was used and the target sample size was 255, determined by identifying the smallest acceptable of a demographic subgroup in which in this situation our UKM medical faculty population size is 700 with a ±5% margin of error and a confidence level of 95% [20]. Eventually, there were a total of 302 respondents, consisting of 150 and 152 Year 1 to Year 5 UKM undergraduate medical students that participated in this study for before and during COVID-19 pandemic period, respectively.

### Materials

Our tool was adapted by Students Motivation Towards Science Learning (SMTSL) survey [21], which has been developed and validated by researchers at the National Changhua University of Education, Taiwan (Cronbach’s Alpha; □ =0.84). It was used to assess the learning motivation of respondents. It consisted of six scales: self-efficacy, active learning strategies, medical learning value, performance goal, achievement goal, and learning environment stimulation. A five-point Likert-type scale was used, and respondents were asked to rate their agreement for each statement as follows: 1=strongly disagree, 2=disagree, 3=no opinion, 4=agree, and 5=strongly agree. Out of a total of 35 questions, there were nine that were reverse items (Question 2, 4, 5, 6, 7, 21, 22, 23, 24). The questionnaire has been modified for medical students and minor adjustments in grammar were made to avoid confusion among the medical students (S1 Appendix). A pilot study was done on 35 UKM medical students (Cronbach’s Alpha; □ =0.91). Demographic data and education background of samples were collected including age of the respondents, year of study, and phase of study. Respondents were also required to rank their preferred choices of digital learning source and choose their frequency of digital learning usage. Feedback and reflections regarding digital learning and learning motivation were also collated from each participant to justify the current situation.

### Procedure

After obtaining Universiti Kebangsaan Malaysia (UKM) Ethics approval (FF-2020-037), a set of questionnaires including information sheet and consent form were distributed via Google Forms through social media platform, WhatsApp. There was no time limit for survey completion and the scores for each of the scales were calculated. Results were recorded using Statistical Package for Social Science (SPSS)

Version 22 by the IBM Corporation, New York, United States. The statistical significance level was set at p<0.05. A summary of social demographic, education background variables, and frequency of digital learning usage was done using descriptive analysis of frequency. The relationship between total score as dependent variable and education background as independent variables was analyzed using cross tabulation analysis. All the variables were transformed by labelling into categorical variables. Descriptive analyses were included for frequencies and percentages of digital learning usage while Student’s t-test and Chi-Square analysis were utilized to determine the difference between groups for selected variables.

## RESULTS

### Demographic Characteristics

A total of 302 Year 1 to Year 5 UKM undergraduate medical students participated in this study, with 150 (49.7%) respondents participating during pre-COVID-19 and the remaining 152 (50.3%) respondents involved during the COVID-19 period. Out of the total, the mean age was 21.94±1.63, and 187 (61.9%) were in the clinical phase of study. Other demographic characteristics are detailed in Table 1.

**Table 1.** Demographic Characteristics of Respondents (n=302).

| Demographic Data |  | Number of Students (%) |
| --- | --- | --- |
| Year of Study | 1 | 61 (20.2) |
|  | 2 | 54 (17.9) |
|  | 3 | 54 (17.9) |
|  | 4 | 71 (23.5) |
|  | 5 | 62 (20.5) |
| Phase of Study | Preclinical | 115 (38.1) |
|  | Clinical | 187 (61.9) |

### Analysis of Digital Learning Usage

The frequency of digital learning usage was being divided into 5 groups; does not use, use less than 1 time per week, 1 to 2 times per week, 3 to 4 times and more than 5 times per week. Respondents rated the frequency based on their usage other than regular classes purposes. During the pre-COVID period, most of the respondents (50%) used digital learning more than 5 times per week and there was a small proportion (6%) of respondents never used digital learning. However, during the COVID-19 period, as medical education has fully transformed to entirely online teaching and learning, all respondents have employed digital learning in their studies with an increased number of respondents using more than 5 times per week (58.55%) as compared to 50% prior to the COVID-19 outbreak (Fig 1A). As most of the UKM medical students had an average of 2 to 3 teachings, group discussions or lab sessions per week, hence the frequency of digital learning usage was further divided into high usage (use at least 3 times and above per week) and low usage (use less than 3 times per week). Overall, most of the respondents had high usage before and during COVID-19 period and there was a rise in high usage during COVID-19 period (86.8%) as compared to pre-COVID-19 period (76.7%) (Fig 1B). Difference in digital learning usage between pre-COVID-19 and COVID-19 period was assessed using Chi-Square analysis (Fig 1C) which showed a significant difference (p<0.05) as students during the COVID-19 period had a higher frequency of digital learning usage as compared to prior COVID-19 period. Among all the respondents who use digital learning, e-books were the most preferred choice for the source of digital learning (40.67%) while audiotapes (6.00%) was the least preferred one before the COVID-19 period. However, videos (38.16%) were the most preferred choice during COVID-19 period. A reduction in preference for e-books (36.84%) and online research articles (11.84%) with an increase in preference for videos (38.16%), online courses (21.05%), games (16.45%), simulation software (11.84%) and audiotapes (9.97%) were noted during this pandemic period (Fig 1D).

**Fig 1.**
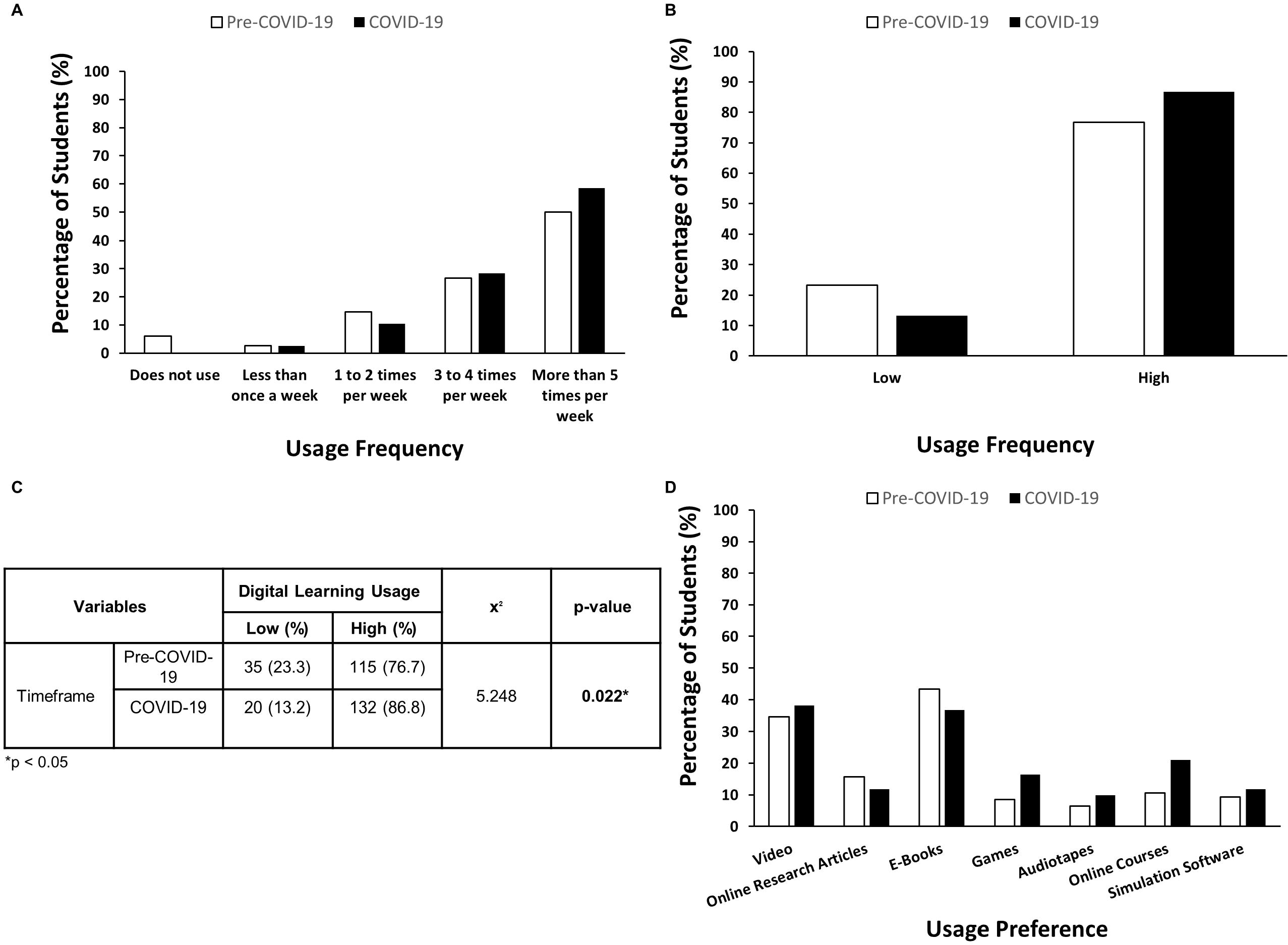
Digital Learning Usage Before and During COVID-19 (n=302).

### Analysis of Learning Motivation

Descriptive statistics were used to provide an overall picture of the learning motivation of this study. Based on the total motivation score, the students can be grouped into 3 levels of motivation; “Low”, “Moderate”, and “High” as previously mentioned [21]. As for prior to COVID-19 period, the samples were normally distributed with an average high learning motivation score of 135.8 ± 11.67 of a total of 175. The mode and median of the total learning motivation score were both 136, while the lowest and highest scores were 102 and 161, respectively. Most of the students showed high motivation (56.9%), followed by moderate motivation (37.9%) and low motivation level (5.2%). On the contrary, during the COVID-19 period, the respondents were also normally distributed with a similar average high learning motivation of 135.59±11.50 with the mode of 132 and 140 and median of 135. However, there was an increment percentage of respondents with high learning motivation (67.80%) (Fig 2A). Difference in level of learning motivation among different variables was analyzed using the Student’s t test. Results showed that the level of learning motivation was significantly different (p<0.01) across the frequency of digital learning usage during the COVID-19 period as higher total scores were obtained with higher frequency of digital learning usage (Fig 2B). Nevertheless, the level of learning motivation between before and during COVID-19 period (Fig 2A), and level of learning motivation across frequency of digital learning usage prior to COVID-19 period showed no significant difference (Fig 2B).

**Fig 2.**
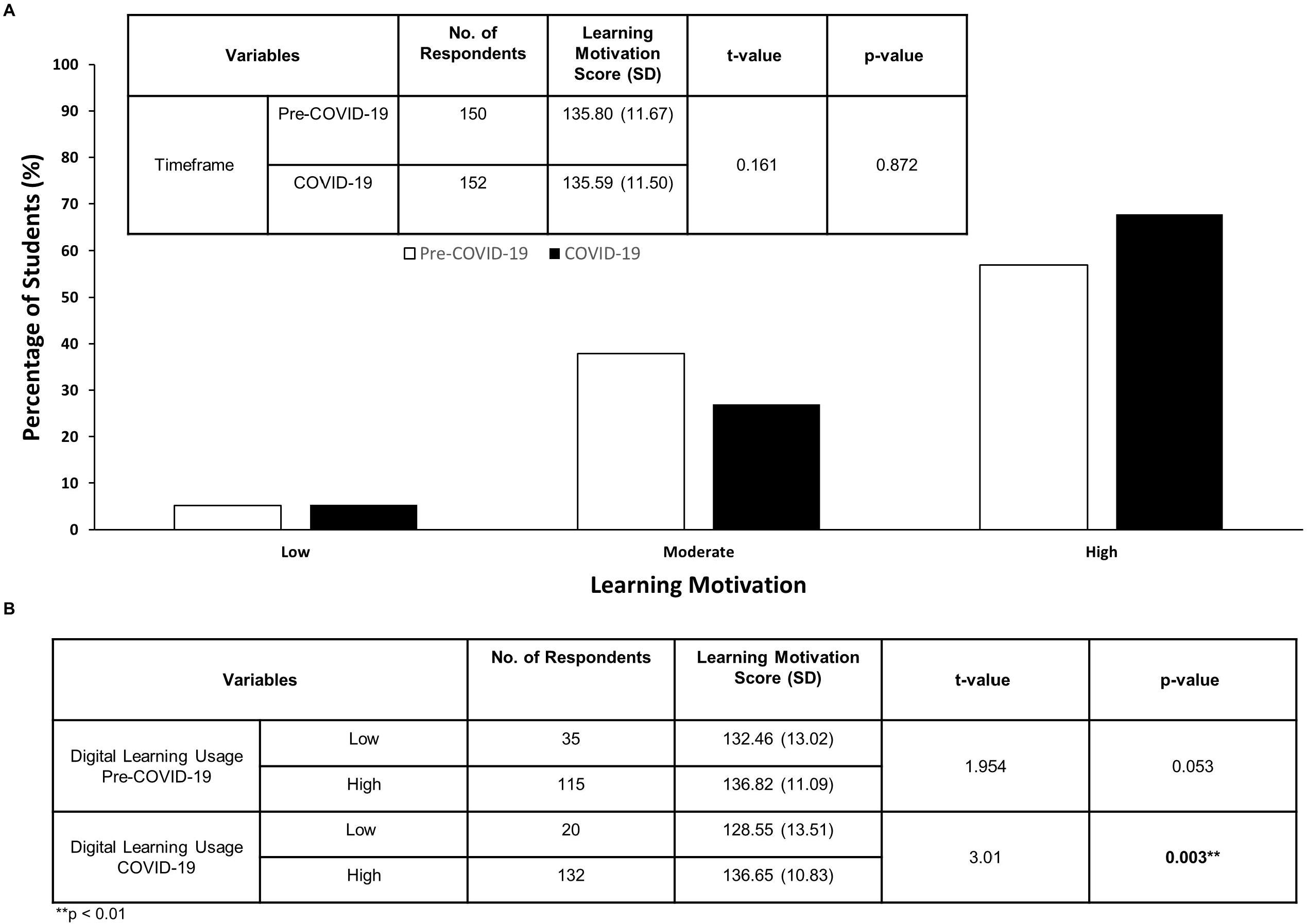
Learning Motivation Before and During COVID-19.

### Analysis of Digital Learning Usage and Learning Motivation among Preclinical and Clinical Medical Students

Difference in frequency of digital learning usage and level of learning motivation before and during COVID-19 period were also assessed among preclinical and clinical medical students respectively via Chi-Square analysis and Student’s t-test. There was a significant difference (p<0.05) in digital learning usage among respondents in the clinical phase during the pre-COVID-19 and COVID-19 period as clinical medical students showed a higher frequency of digital learning usage during the COVID-19 outbreak (87.1%) as compared to before the pandemic period (74.5%) (Fig 3A). However, there was no significant difference in level of learning motivation among clinical medical students before and during COVID-19 period (Fig 3B). No significant difference was noted as well for both digital learning usage and level of learning motivation between pre-COVID-19 and COVID-19 timeframe among preclinical medical students (Figs 3A and 3B).

**Fig 3.**
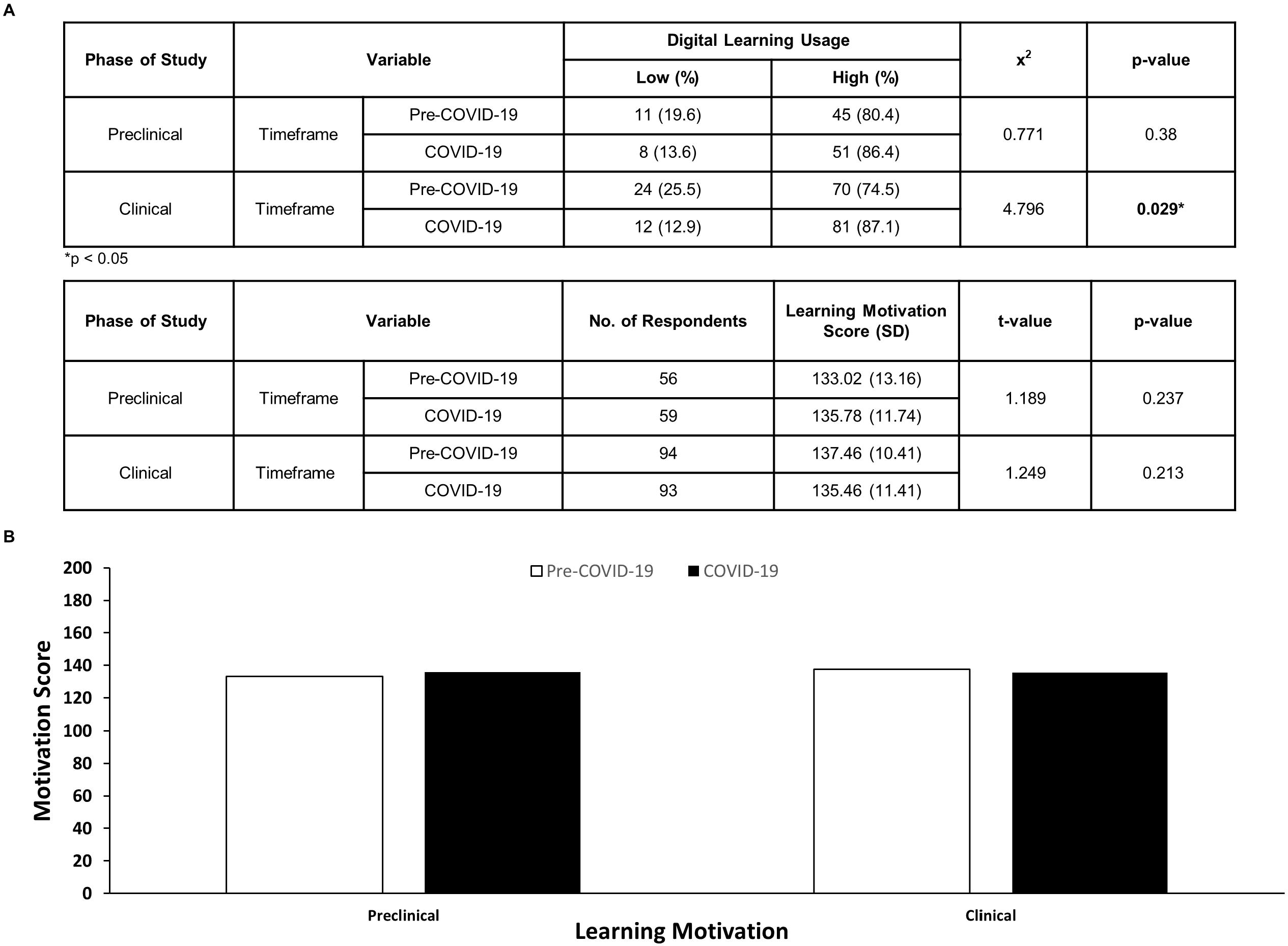
Learning Motivation and Digital Learning Usage Based on Phase of Study.

### Analysis of Themes of Learning Motivation (LM) during COVID-19 Period

We have gathered feedback and reflections and further collated them into 11 emerging themes that featured the learning motivation of UKM medical students during COVID-19 period (Table 2), which were further summarized into 2 main themes; intrinsic and extrinsic motivations (Fig 4). Intrinsic motivation in learning refers to the action of self-learning because of inherent interest or satisfaction, whereas extrinsic learning motivation pertains to learning to attain some other outcomes that are unrelated to the action. We labelled respondents as “R” in our transcripts.

**Table 2.** Quotes from respondents to highlight main themes of learning motivation.

| Quote | Emerging theme | Main theme |
| --- | --- | --- |
| “I set up a to-do list every day and focus on things I want to study in small sessions.” (R2) | Self-motivation | Intrinsic motivation |
| “Regardless whether we're in difficult times, learning is a continuous journey, the one that can only stop you from learning new knowledge is your own self.” (R12) | Self-discipline |  |
| “Very badly. No motivation most of the day” (R29) | Self-adaptation |  |
| “To pass the year and move on” (R139) | Feeling indifferent |  |
| “Weekly teaching and lecture motivate me to understand things better” (R1) | Online teaching | Extrinsic motivation |
| “My supervisor is the main reason why I am able to cope with this kind of learning method.” (R4) | Lecturer |  |
| “I watch videos to understand better.” (R54) | Digital learning using different resources |  |
| “Always remind myself about the pro exam.” (R14) | Examination |  |
| “Ask motivation from friends and family” (R11) | Family |  |
| “By continuously keeping in touch with my friends, making a checklist of the subjects that I need to cover, having a conducive study area, online group study with friends.” (R31) | Friend |  |
| “Not having a conducive studying environment at home ...” (R43) | Environment |  |

**Fig 4.**
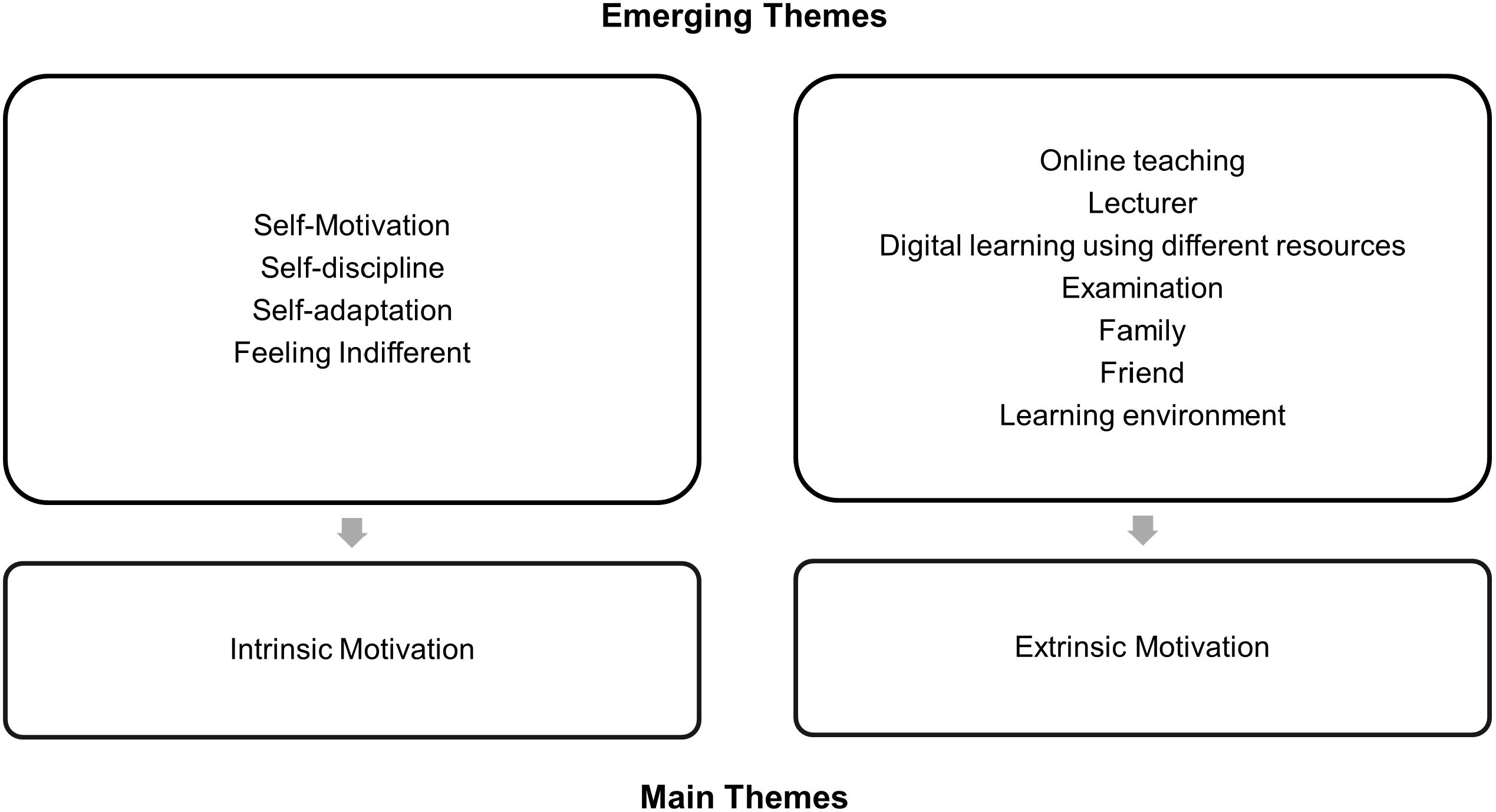
The summary of emerging themes based on the feedback on learning motivation.

#### Theme LM1: Intrinsic motivation

When asked about learning motivation, there were 28 responses that pointed towards self-motivation during COVID-19 pandemic. Most of the responses mentioned about reminding oneself subconsciously about the reason for choosing the medical profession and the necessity of continuous learning.

> “Regardless whether we’re in difficult times, learning is a continuous journey, the one that can only stop you from learning new knowledge is your own self.” (R12)

Some medical students also boosted their self-motivation via other sources available while learning at home.

> “Watch vlogs of medical student like “Ali Abdal” to make me feel like studying.” (R59)
>
> “By watching motivational videos.” (R64)

Another emerging theme that was categorized under intrinsic learning motivation would be self-discipline. More time to study at home does not necessarily mean more effective learning when students are expected to learn on their own. There were 50 responses that portray how medical students apply the principle of self-discipline as learning motivation.

> “I set up a to-do list every day and focus on things I want to study in small sessions.” (R2)
>
> “A lot of self-study, researching materials and discussions with lectures.” (R45)
>
> “Study a lot and never forget to rest in between online sessions.” (R110)

Also, there were 3 responses that mentioned the feeling of indifference despite the transformation.

> “To pass the year and move on.” (R39)

Shifting from face-to face learning to digital learning is an enormous challenge to medical students especially those clinical students who used to practice clinical skills in hospital settings. This transformation requires the skill of self-adaptation in oneself. However, despite various themes of learning motivation attained, there were 4 responses that point towards incapability to adapt due to lack of both intrinsic and extrinsic motivation. 3 out 4 responses were from clinical students.

For intrinsic motivation, one student said, “Very badly. No motivation most of the day.” (R30) and another student quoted, “Difficult.” (R55)

#### Theme LM2: Extrinsic motivation

Learning environment was the extrinsic motivation that contributed to the inability to adapt as quoted by one student,

> “…Not having a conducive studying environment at home and also a lot of reading materials have been left at the college…” (R43)

As online classes have become the bread and butter routine of everyday life, there were students (n=10) who said that online learning motivated them to study in order to understand the lessons better.

> “Weekly teaching and lecture motivate me to understand things better” (R1)

Among the students whose learning motivation was extrinsic, 6 of them wrote lecturers can be a source of learning motivation.

> “My supervisor is the main reason why I am able to cope with this kind of learning method.” (R4)

Different sources of digital learning which were found online such as research articles, ebooks, and educational videos were considered as extrinsic motivation because these sources were very informative and easily accessible.

> “I watch videos to understand better.” (R54)

Cancellations or delays of examinations over the COVID-19 pandemic do not demotivate the students in terms of learning persistently at home, in contrast, upcoming delayed examinations have become a learning motivation to 18 students, especially those who are in final year.

> “Counting days for professional exams.” (R7)

A total of 32 students think that family (n=6) and friends (n=26) can act as a strong support for them to study at home during COVID-19.

> For example, one remark was, “Ask motivation from friends and family.” (R11)
>
> Another student said, “My friends and I have discussions frequently regarding questions we encounter during our teachings…” (R26)

### Analysis of Reflections of Methods (RM) to Improve Digital Learning

Feedback on digital learning improvement were collated (Table 3), consisting of a total of 18 emerging themes, which were further summarized into 5 main themes (Fig 5).

**Table 3.**
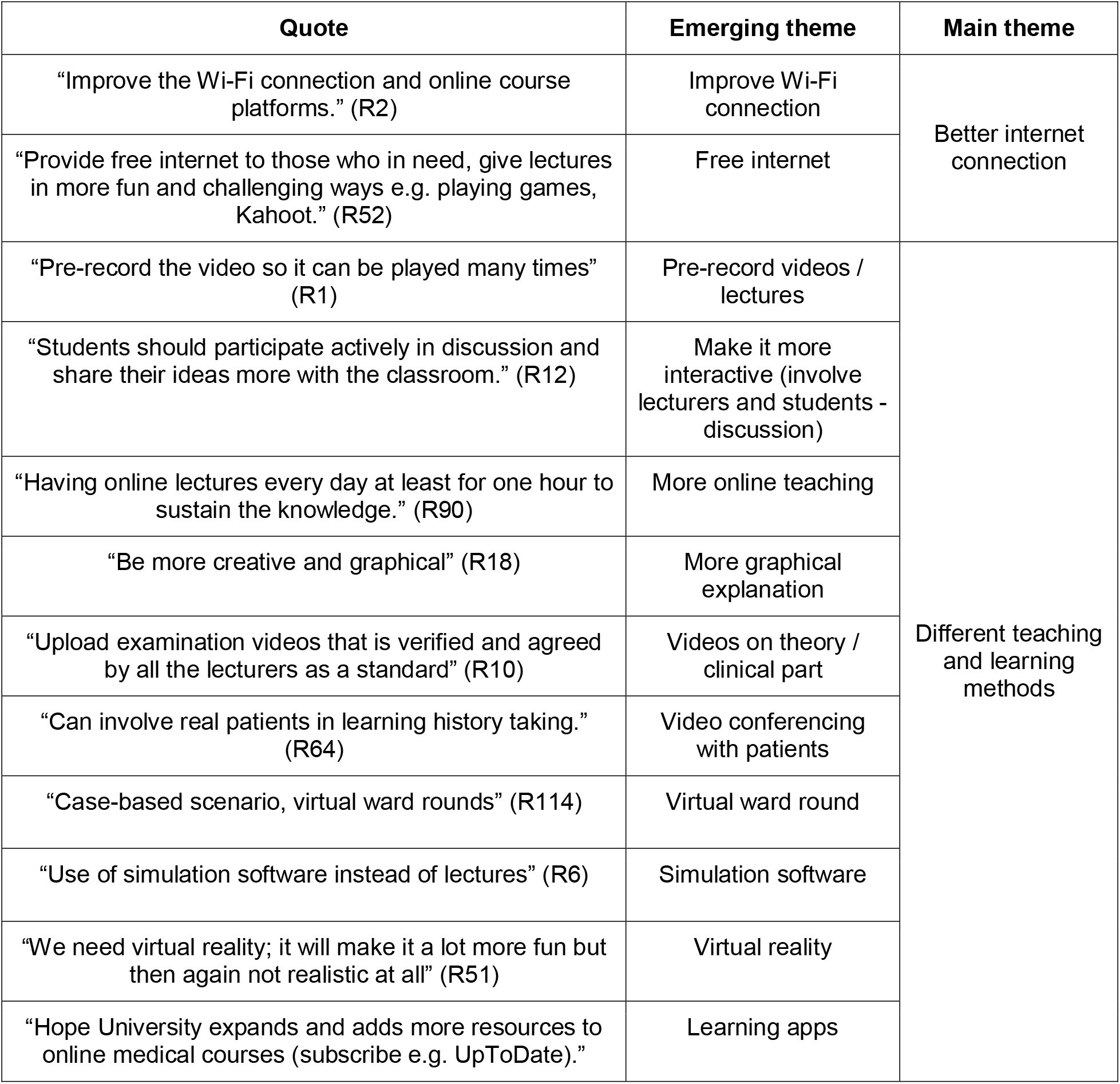
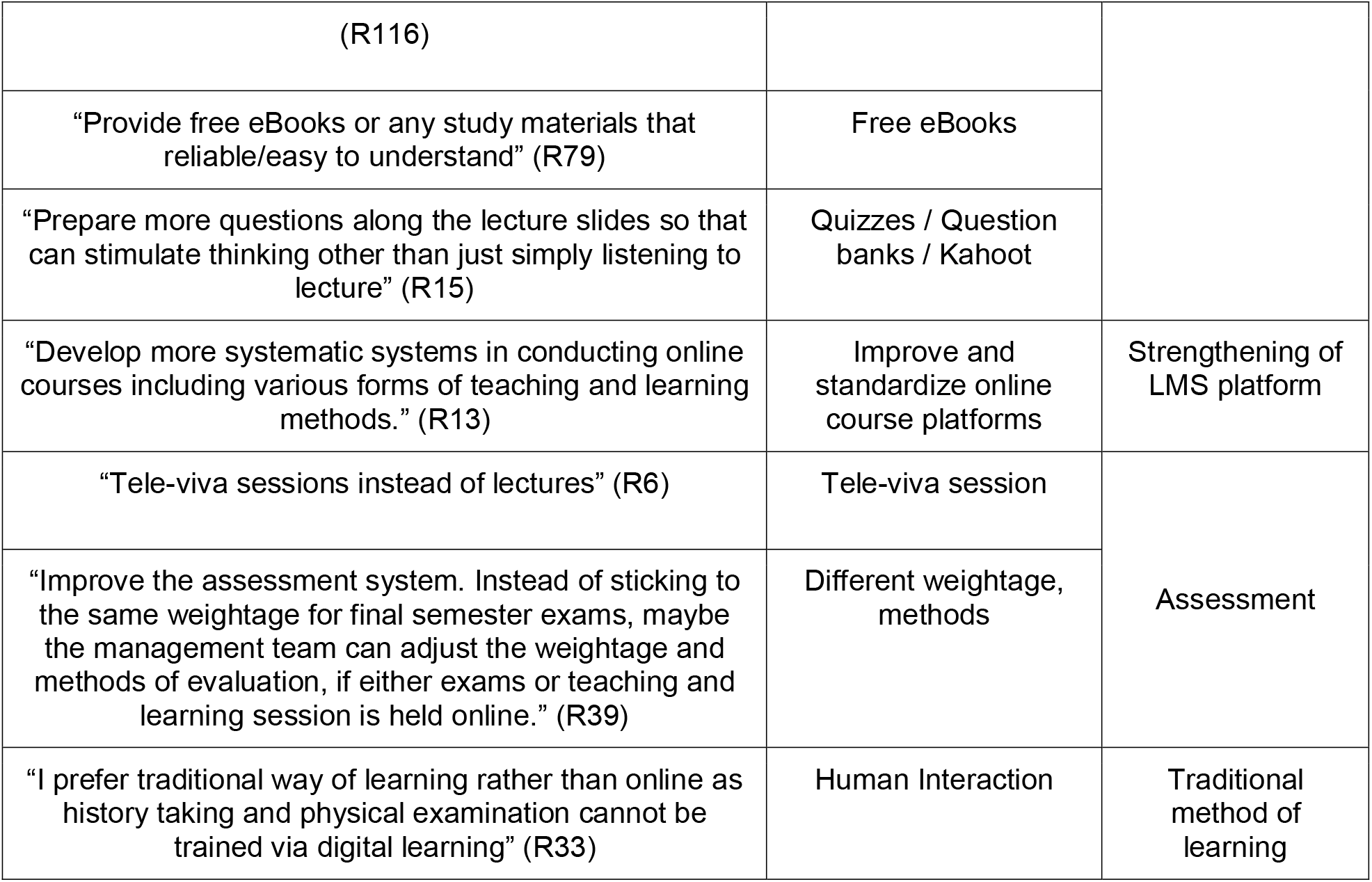
Quotes from respondents to highlight main themes of digital learning improvement.

**Fig 5.**
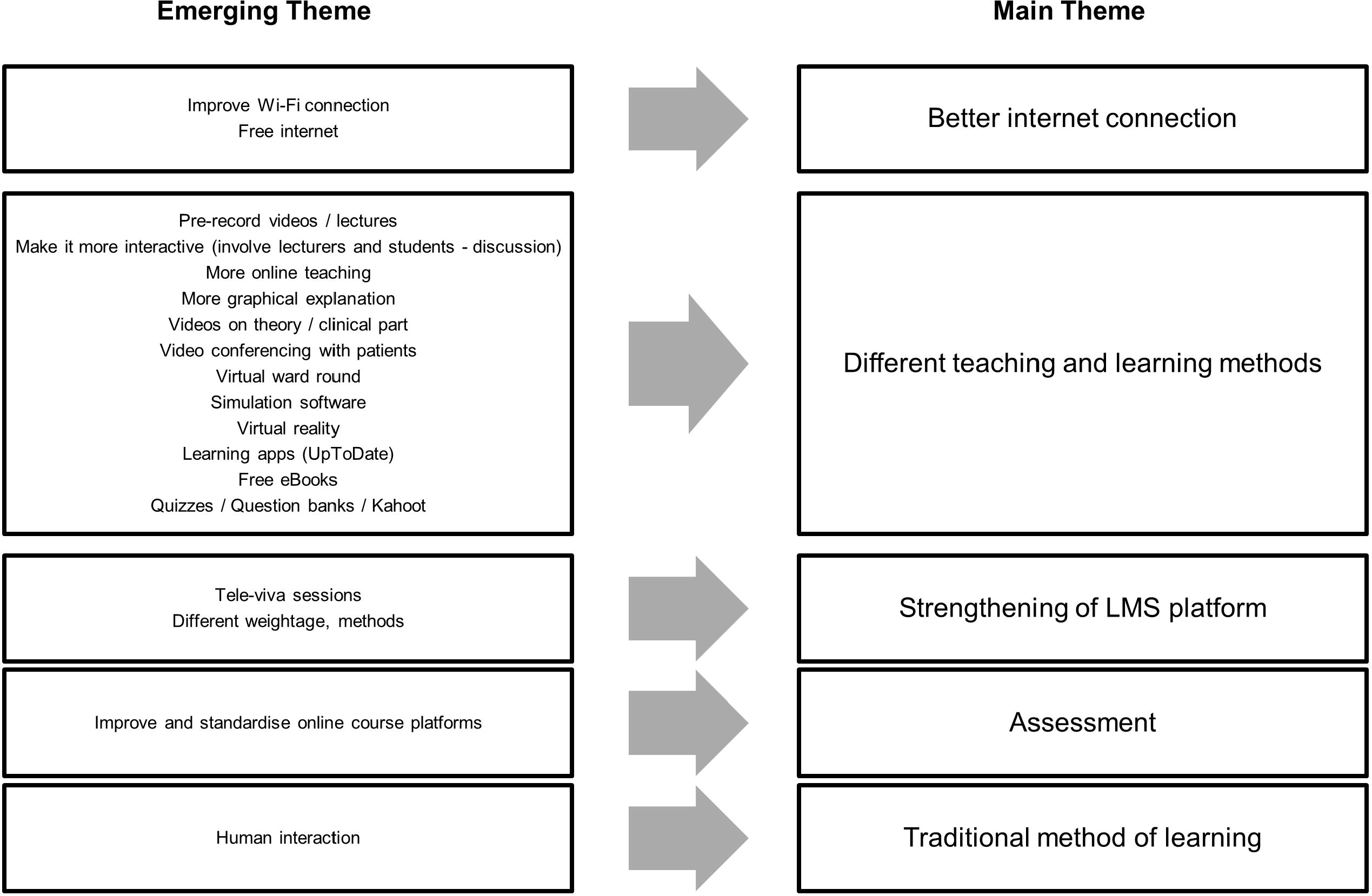
The summary of emerging themes based on the feedback on digital learning improvement.

#### Theme RM1: Better internet connection

Online learning is very dependent on internet access, without a stable internet connection, the learning process would be interrupted since most of the materials and resources were only available online. In this theme, there were two groups of responses which included improvement of Wi-Fi connection (16 responses) and free internet access (2 responses).

> “Improve the Wi-Fi connection and online course platforms.” (R2)
>
> “Provide free internet to those who need it, give lectures in more fun and challenging ways e.g. playing games, Kahoot.” (R52)

#### Theme RM2: Different teaching and learning methods

Twelve different approaches in medical learning were collated from the reflection under the main theme of different learning methods. Due to occasional unstable Internet connection, some students were unable to access synchronized online classes, there were nine students suggested that lectures and educational videos to be pre-recorded before class or/and recorded during the class as a reference.

> “Record teaching video for practical besides concept lecture because it is better than online discussion. Sometimes the line (internet) does not cooperate, I can’t get the full content of the discussion. “(R32)

There were students (n=27) who deemed e-learning as a more interactive way of learning and involved inter-communication between both lecturers and students instead of a didactic way of teaching.

> “More group discussions and problem-based learning than mass lecture.” (R46)

While all medical students were staying at home under Movement Control Order, three students mentioned that online teaching should be delivered more frequently, and one of them give an example,

> “Having an online lecture every day at least for one hour to sustain the knowledge.” (R90)

To enhance understanding on certain topics, especially lessons involving anatomy and physiology, there were two comments about preparation of more graphical explanation, for instance, lecturers may incorporate more mind maps or diagrams when delivering lectures.

> “More exciting and easier to understand by using more pictures along with the explanation.” (R144)

For substitution of physical examination, 22 responses suggested that it can be learned through demonstration via video and explanation of each step can be recorded simultaneously.

> “Upload (clinical) examination videos that are verified and agreed by all the lecturers…” (R10)

History taking can be carried out by doing video conferencing with patients whose condition was stable to be interviewed as suggested by two students.

> “Can involve real patients in learning history taking.” (R64)

There was one response that commented about virtual ward rounds to replace the routine ward round done in a hospital setting.

> “…virtual ward rounds” (R114)

Learning via simulation software was also mentioned by 23 students.

> “Can improve session like “DxR Clinician” so student can improve (on) their clinical reasoning skills” (R14)

Three responses suggested virtual reality to replace the clinical exposure learning for medical students.

> “We need virtual reality; it will make it a lot more fun but then again not realistic at all” (R51)

There were two students who wished to subscribe to more learning apps during this COVID-19 period.

> “…. subscribe to UpToDate” (R21)

For students who used to borrow medical books from the library or left their study materials at the students’ accommodation, there was one suggestion about free e-books which can be accessed online.

> “… provide free eBooks or any study materials that are reliable/easy to understand…” (R79)

To consolidate one’s knowledge and explore their own understanding on certain topics, there were 16 responses mentioned about quizzes, question banks or Kahoot.

> “…provide practice questions / clinical scenarios as homework, to be discussed further online” (R21)

#### Theme RM3: Strengthening of LMS platform

Learning Management System (LMS) is another crucial component in transformations of medical education via digital learning. An ideal LMS will provide a standardized platform to all students to access e-learning contents systematically, interact with each other while lecturers are able to track progression of students. Some medical students recommended that UKM’s LMS, UKMFolio™ should be improved as shown below,

> “…my suggestion is that our university should prepare a dedicated website for online learning where all the syllabus is structured.” (R66)
>
> “Standardizing the way, the instructions are given such as displaying the submission date and time in a column where all students and lecturers can see clearly.” (R74)
>
> “(LMS) a stable platform where students and lecturers can communicate well because it’s more like one-way communication now. And taking longer than usual.” (R82)

#### Theme RM4: Assessment

A small proportion of students (n=2) suggested that the assessment method should be modified to be in line with the modifications upon implementation of fully digital learning.

> “Improve the assessment system. Instead of sticking to the same weightage for final semester exams, maybe the management team can adjust the weightage and methods of evaluation, if either exams or teaching and learning session is held online.” (R39)

#### Theme RM5: Traditional method of learning

A total of four students insisted on having traditional methods of learning as they believed that clinical components especially cannot be replaced with digital learning. 3 out of these four students were clinical students. 2 of them even proposed that they were willing to postpone the current semester and graduate at a later date until they were being allowed to enter the hospital and resume the usual way of learning from patients.

> “I prefer traditional ways of learning rather than online as history taking and physical examination cannot be trained via digital learning.” (R7)
>
> “Nothing, no online classes ever can beat the actual learning method by seeing patients. Postpone the semester.” (R130)

## Discussion

Conventional medical education pedagogies have been emphasizing on clinical internships involving direct interaction with patients while digital learning only plays a complementary role. Digital learning has drawn a great deal of attention in this 21st century due to its flexibility, deliverability, freedom, and independence of learning. However, it is generally believed that digital learning could not replace the traditional education methods [22] as the nature of the course of medicine involves substantial hands-on skills which digital learning lacks. Indeed, the sudden implementation of fully online medical education upon introduction of Movement Restriction Order (MCO) has left all faculty members unprepared with the unforeseen changes in the learning methodology, which actual outcome is yet to be observed. One of the main concerns is internet connection issues such as poor internet coverage, low internet speed, and network congestion. This has been the main challenge faced by UKM undergraduate students throughout their studies [23]. In fact, 40% of our medical students are having poor internet connection (<5Mbps). Feedback from the medical students also showed that internet connection must be improved, and free internet should be provided in order to achieve efficient digital learning. Indeed, the initiative of allocating free 40GB data connection by UKM and YTL Communications to each UKM student during this pandemic period should be applauded and continued to avoid infrastructure issues from deterring digital learning in the future. On the other hand, learning motivation, specifically autonomous motivation has been playing a crucial role in academic success and lifelong learning in the career pathway as a medical practitioner. At the time of writing, our study is the first research that explores the relationship between digital learning and learning motivation among medical students during the COVID-19 pandemic as compared to the period prior to the pandemic in the Malaysian setting.

Generally, the usage of digital learning before and during COVID-19 is high with a significantly higher digital learning usage during the outbreak period. This is reasonable as the transformation into fully digital learning upon enactment of MCO has left medical students with no choices but to rely on digital learning. Multimodal has been the preference style of learning especially among those universities that employ integrated curriculum, which is similar to UKM as the core component of curriculum generally focuses on student based learning, mainly discussions, problem based learning and practical instead of the usual didactic learning [24-26]. Although medical students use all styles of learning, the majority use aural method [27-30] with aural kinesthetic being the most common combination of learning style [30] while reading or writing [27, 28] is the least preferred style in the Visual, Auditory, Reading or Writing and Kinesthetic (VARK) model. Indeed, we believe that this medical education transformation has caused certain changes and impacts on learning styles among medical students. Lecturers and medical students must make use of the available resources in digital learning to get the utmost knowledge and skills for practical clinical components that was initially obtained through clinical clerkships at the hospital. This was in accordance with our results as we can see that the percentages of student’s preference shifted towards the usage of videos, online courses, games, simulation software and audio recording during the outbreak period. Suggestions from our medical students also showed that during the outbreak, they hoped that they could be provided with more interactive forms of digital learning, simulation software, videos regarding the topics to be learnt and recordings of lectures, which are all in line with the preferred source of digital learning. This is reasonable as all these sources are either related to aural style of learning or it can partially replace kinesthetic style since students do not have any direct experience with patients via clerkships in hospitals during the pandemic period. Indeed, medical students have positive perception towards the use of simulation based learning [31] and virtual reality [32] as they provide hands on practice and improve learning competency with clinical reasoning skills especially when there is limited availability of patients in hospitals. With the availability of simulation labs in UKM, students should be encouraged to make use of it to gain medical-procedural experience while more training workshops should be provided for lecturers so that it could be used as an additional tool for medical teaching and learning.

Our study showed that most medical students possessed high learning motivation in using digital platforms as a process of learning. This trend was found similar before and during COVID-19. There was no significant difference in level of learning motivation within this duration even though digital learning usage was significantly higher during the outbreak period. This shows that learning motivation among medical students is not directly affected by digital learning usage but instead there are other factors playing a role in driving continuous motivation. The most plausible factor could be autonomous motivation as medical students are known to have high intrinsic motivation as compared to undergraduate students of other courses [33]. High motivation has been one of the vital components during selections or interviews for the course of medicine [34-36]. Previously, it has also been reported that there was no significant difference in the level of intrinsic motivation among medical students who participated in high-fidelity mannequin simulation, as the medical students were found to be originally highly intrinsically motivated [37]. This was well reflected by our medical students who portrayed intrinsic motivation, mainly self-motivation, and self-discipline in coping with this tough time. For instance, some of them will motivate themselves to keep on learning by recalling the actual intention of the onset of medical pursuit while some laid out an organized study plan with daily learning objectives and they would adhere to it to ensure that they learn something every day. However, the significantly higher learning motivation with high digital learning usage during the pandemic period could be explained as online learning being the only source of learning during COVID-19. Thus, medical students with pre-existing high intrinsic motivation will use all available resources for their study and lean towards digital sources.

An in-depth review on the students based on two phases of study; preclinical and clinical, has shown that there was no significant difference in digital usage and learning motivation for the preclinical students in both pre-COVID-19 and COVID-19 periods. Similar trend was noted among clinical students despite significantly higher digital learning usage. This is probably due to the curriculum structure of the medical program in UKM, as the preclinical phase mainly consists of theoretical studies with problem-based learning which may ease the transition into digital learning as compared to clinical phase that mainly involves hospital clerkships with direct interaction with patients. As compared to preclinical students, greater responsibility for patients and clinical practice drives motivation for clinical students that reinforces and sustains the students’ motivation profile [38, 39]. Thus, clinical students would be more impacted as they have missed the most crucial part of learning directly from patients, in which up to recently, there is a lack of data that this could be replaced with digital learning. In fact, there was feedback from the students insisting that the clinical components must be taught in hospitals and can never be replaced by digital learning. Studies have also shown that a conducive learning environment plays a crucial role as it correlates with a good motivation [40, 41]. Our results showed that the learning motivation among clinical students is slightly lower during the COVID-19 period as compared to the usual times. This may be due to the sudden change in learning environment and methodology thus the need to adapt with such changes. Reflections from our medical students stated that the unconducive learning environment at home was a discouraging factor. This could be due to inaccessibility of certain learning materials such as online research articles or certain online learning platforms that are only accessible at the university area. Hence, we propose that more online learning platforms like UpToDate and full text journals should be subscribed by the university and more importantly, these should be accessible without any time and place restrictions. Besides, as suggested by our students, an ideal LMS that enables students to access all learning materials while encouraging discussions between lecturers and students via online assessments such as quizzes and viva sessions are required if the university were to embark on a successful digital learning experience.

This study was conducted with a few potential limitations. Since this was a cross-sectional study, we could not accurately elicit the direct causal relationship between digital learning and learning motivation among medical students. Additional longitudinal studies are recommended in the future to further explore the relationship between digital learning and learning motivation and feasibly on students’ learning performance. Next, since only UKM medical students are involved in this study, there are limitations to the representativeness and generalizability of the findings to another setting in other medical faculties. Additionally, the likelihood of respondents giving socially desirable responses as participants may answer the questionnaire positively based on what they perceive to be expected of them since our study used self-reported data.

## Conclusion

The usage of digital learning is established among UKM undergraduate medical students with e-books as the choice of source prior to the COVID-19 pandemic. However, over the course of the COVID-19 outbreak, there was a significantly higher digital learning usage among the students with videos being the preferred source. Despite the sudden increment in digital usage, there was no significant difference in learning motivation between the two time periods. A review on the phase of study showed that there was only a significantly higher digital learning usage among clinical students. It is also shown that the students generally had higher intrinsic motivation as compared to extrinsic motivation with self-motivation being the leading theme. Thus, medical educators could focus on extrinsic motivation by the utilization of digital learning as an additional driving factor to the students as the students already have high intrinsic motivation as compared to extrinsic motivation. This could be enhanced by investing in necessary state-of-the-art innovation of online teaching and learning methods to transform medical education with supplementation of high-end technology. This could be a platform for medical students to learn to be independent in learning new information in the vast pool of digital information and to apply life-long learning in medical practice.

## Data Availability

All data referred to in the manuscript is available as a supplementary file.

https://drive.google.com/drive/u/0/folders/1-R06hPBEO9a8fp8BYwocoz7sBlmBfm25

## Acknowledgement

We would like to thank Associate Professor Dr. Azimatun Noor Aizuddin for guiding us through the process of statistical analysis. Lastly, we would like to thank the medical students currently enrolled in the 2019/2020 session who actively participated in this study.

## Supporting information

**S1 File. Medical Students Learning Motivation Survey.**

**S2 File. Raw Data Pre-COVID-19**

**S3 FIle. Raw Data COVID-19**

## Notes

### Competing Interest Statement

The authors have declared no competing interest.

### Funding Statement

This study has been funded by Faculty of Medicine, UKM (FF-2020-037) to JXL, AHAA, JYN, and NASI. The funder had no role in study design, data collection and analysis, decision to publish, or preparation of the manuscript.

### Author Declarations

This study has been reviewed and approved by the Research Ethics Committee, The National University of Malaysia (UKM) UKM/PPI/111//8/JEP-2019-702

